# METABOLOMIC INSIGHTS: LC-MS PROFILING OF HUMAN PLACENTAL TISSUE FROM SSRI-TREATED PREGNANCIES

**DOI:** 10.1101/2025.05.12.25327429

**Authors:** Anna Itkonen, Olli Kärkkäinen, Marko Lehtonen, Heidi Sahlman, Leea Keski-Nisula, Jaana Rysä

## Abstract

Antenatal depression, a common pregnancy complication, poses significant risks if untreated. Consequently, pregnant mothers are prescribed selective serotonin reuptake inhibitors (SSRIs). Based on their pharmacological action, SSRIs have the potential to disrupt serotonin signaling, which is crucial for placental function and fetal development. SSRIs are associated with adverse effects on both placenta and fetus. Understanding placental metabolism is essential for assessing fetal exposure to SSRIs. However, the impact of maternal SSRI usage on placental metabolism remains understudied.

We performed a comprehensive determination of the placental metabolome by nontargeted liquid chromatography-mass spectrometry metabolomics approach to determine whether SSRIs alter placental metabolic functions. A total of 48 placental samples from individuals using SSRI medication throughout the pregnancy (n = 24) and non-depressive controls without antidepressant medication (n = 24) were included.

We observed significant alterations in placental metabolic profiles among individuals using SSRIs, potentially indicating a response to changes in placental redox homeostasis and energy metabolism. Furthermore, four of the altered metabolites were positively correlated with the 1- and 5-minute Apgar score in SSRI-treated pregnancies, indicating that higher metabolite levels may correlate with better birth outcomes. Given the limited research on placental metabolomics, our exploratory study provides new insights into SSRI-induced changes in the placenta.

## INTRODUCTION

Depression is a common complication during pregnancy, with untreated antenatal depression posing significant risks to both the mother and the developing fetus [1]. Emerging evidence from both rodent and human studies suggests that the placenta can transmit maternal depression and stress to the fetus, potentially resulting in long-term mood disorders in offspring [2]. Thus, selective serotonin reuptake inhibitors (SSRIs) are frequently prescribed during pregnancy.

SSRIs exert their effects by inhibiting serotonin (5-HT) reuptake through serotonin transporters (SERT), increasing extracellular 5-HT levels in the central nervous system and peripheral tissues, including the placenta [3]. In the placenta, SERT is expressed on the maternal-facing brush border membranes of the trophoblast cells [4]. Furthermore, placenta exhibits a 5-HT system, which is crucial for pregnancy progression and early embryonic development, regulating cell division, morphogenesis, and neuronal plasticity and supporting placental blood flow and fetal brain development [5–7]. Thus, the placenta is vulnerable to disruptions in placental 5-HT homeostasis. By inhibiting placental SERT, SSRIs may interfere with 5-HT signaling, critical for placental function and the development of the fetal central nervous system and other organs [6,8]. SSRI usage during pregnancy is associated with increased risks of major congenital malformations, congenital heart defects, preterm birth, neonatal adaptation symptoms, and persistent pulmonary hypertension of the newborn, particularly with paroxetine and fluoxetine [9].

Reported by us and others, SSRIs have the potential to alter placental functions critical for fetal development such as gene expression, and the function of transporters, and enzymes [3,10–12]. Theoretically, these impairments could be reflected in the placental metabolome. Earlier metabolomic studies link SSRIs with circulating metabolic disturbances. In non-pregnant populations, SSRI use has been linked to changes in glucose metabolism, lipid profiles, and weight gain [13]. These systemic metabolic changes suggest that SSRIs may disrupt not only neural signaling but also broader physiological pathways that are essential for cellular homeostasis.

Despite the observed effects of SSRIs on the circulating metabolome and placental function, there is a notable lack of data assessing how SSRIs affect the placental metabolome. Since the placenta acts as the key intermediary between the mother and the developing fetus, understanding its metabolic response to SSRI exposure is critical for elucidating potential risks posed to the fetus. To address this gap in knowledge, we conducted a comprehensive metabolic profiling of placental tissues from SSRI-treated pregnancies using a nontargeted liquid chromatography-mass spectrometry (LC-MS) approach. LC-MS offers the broadest scope of metabolome analysis among available methods, capable of elucidating metabolic pathway regulation and diverse exposures [14]. Our study aims to characterize the differences in the placental metabolome between SSRI-treated pregnancies and control pregnancies without antidepressant treatment to better understand the placental responses of SSRIs. Additionally, we sought to explore the association between placental metabolic profiles and neonatal and maternal outcomes, such as birth weight, Apgar scores, and placental weight.

## RESULTS

### Clinical data

The clinical characteristics of the cohort exhibited a uniform distribution, although SSRI-cases had significantly higher placental weights (Table 1). The most often used SSRI was citalopram, accounting for 50% (n=12) for the cases, followed by escitalopram (25.0%, n=6), sertraline (20.8%, n=5), and fluoxetine (4.2%, n=1) (Supplementary Table 1). Depression was the most frequent indication for the SSRI treatment (62.5%, n=15), followed by panic disorder (20.8%, n=5), anxiety (8.3%, n=2), bipolar disorder (4.2%, n=1) and muscle spasm (4.2%, n=1).

**Table 1.** Clinical characteristics of study participants with selective serotonin reuptake inhibitor users (SSRI) (n = 24) and controls without antidepressant medication (n = 24)

| Variable | Control | SSRI | p-value* |
| --- | --- | --- | --- |
|  | n = 24 | n = 24 |  |
| Age, years | 29.0 ± 4.2 | 30.8 ± 4.8 | 0.179 |
| Parity | 0.87 ± 1.0 | 1.12 ± 1.0 | 0.343 |
| Self-reported smoking |  |  |  |
| Not smoking | 13 (56.5) | 19 (86.4) |  |
| Quit smoking before pregnancy | 3 (13.0) | 0 (0.0) | 0.994<sup>a</sup> |
| Smoked during pregnancy | 7 (30.4) | 3 (13.6) | 0.115<sup>a</sup> |
| Cotinine, ng/ml<sup>1</sup> | 3.92 ± 16.3 | 4.89 ± 13.7 | 0.177 |
| Co-medication | 9 (37.5) | 15 (62.5) | 0.083 |
| BMI at the beginning of pregnancy, kg/m<sup>2</sup> | 24.4 ± 4.1 | 28.2 ± 8.7 | 0.063 |
| BMI before delivery, kg/m<sup>2</sup> | 29.1 ± 4.3 | 32.7 ± 8.0 | 0.056 |
| Number of fetuses | 1.0 ± 0.0 | 1.0 ± 0.2 |  |
| Gestational diabetes mellitus<sup>2</sup> | 4 (16.7) | 6 (25.0) | 0.725 |
| Preeclampsia<sup>2</sup> | 1 (3.4) | 2 (4.0) | 1.000 |
| **Obstetric and early neonatal characteristics** |  |  |  |
| Gestational weeks at delivery |  |  | 1.000 |
| Term ≥ 37 | 23 (95.8) | 23 (95.8) |  |
| Moderately preterm 32-36 | 1 (4.2) | 1 (4.2) |  |
| Mode of delivery<sup>3</sup> |  |  | 0.187 |
| Vaginal | 21 (95.5) | 18 (78.3) |  |
| Caesarean section | 1 (4.5) | 5 (21.7) |  |
| Placental weight, grams<sup>2</sup> | 548.9 ± 86.1 | 646.7 ± 124.7 | **0.040** |
| Birth weight, grams | 3364 ± 455 | 3526 ± 561 | 0.592 |
| Placental weight / Birth weight -ratio | 15.71 (0.04) | 17.83 (0.06) | 0.154 |
| Infant sex (n male) | 10 (41.7) | 16 (66.7) | 0.147 |
| Apgar scores at 1min | 8.79 ± 0.7 | 8.33 ± 1.8 | 0.635 |
| Apgar scores at 5min | 9.16 ± 0.5 | 8.71 ± 1.1 | 0.113 |

Data is shown as mean ± SD or n (%). \* Welch's t-test or Wilcoxon rank-sum test for continuous variables, Fisher's exact test for categorical variables. Logistic regression <sup>a</sup> the values were correlated to "not smoking". <sup>1</sup> n = 47, <sup>2</sup> n = 46, <sup>3</sup> n = 45, BMI; Body mass index, Co-medication; reported use of other medication than selective serotonin reuptake inhibitors

### Metabolite profiling

In the metabolomics analysis, we measured a total of 18162 molecular features (Supplementary Table 2), of which 3086 features had raw p-values <0.05 and 862 features had false discovery rate (FDR)-corrected q-values <0.05 (Supplementary Table 3 shows features with MS/MS data). Principal component analysis (PCA) showed some separation between the SSRI-treated cases and controls based on the first two principal components (Figure 1A). The results of the PLS-DA model with two components (R2Y(cum)=0.96, Q2(cum)=0.62) were in line with the results from the feature-wise analyses (Figure 1B). The identified metabolites with p-value <0.05 are depicted in Figure 1C.

**Figure 1.**
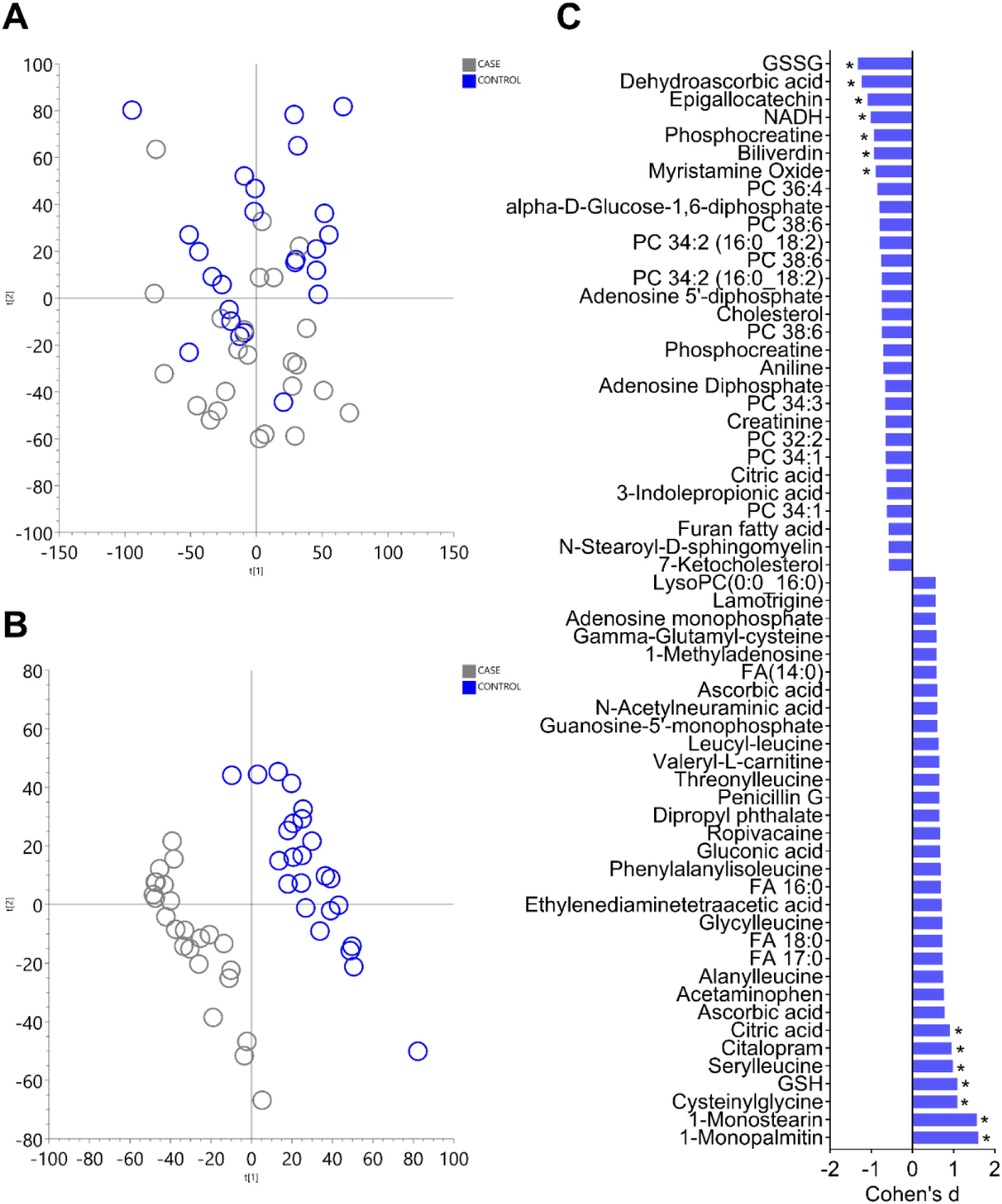
Overview of the metabolite profiling results from the placental samples. Panel A, principal component analysis (PCA) score plot showing first two components explaining 9% and 8% of the variation in the data are shown. The second component shows some separation of the selective serotonin reuptake inhibitor (SSRI) -cases (n=24) and controls (n=24). Panel B, partial least sum of squares discriminant analysis (PLS-DA) score plot of metabolomic profiles of SSRI-cases compared to controls (2 components, R2Y(cum)=0.96, Q2(cum)=0.62). Panel C shows all the identified metabolites with p-value below 0.05 and Cohen’s d effects sizes. A positive Cohen’s d value indicates that the metabolite abundance was increased in SSRI-cases, and a negative value indicates a decrease in SSRI-cases compared to controls. Metabolites with q-value below 0.05 and VIP-values (from PLS-DA) above 1.5 are marked with asterisk (*). GSSG, oxidized glutathione; NADH, nicotinamide adenine dinucleotide; PC, phosphatidylcholine; FA, fatty acid; GSH, reduced glutathione.

Notably, after correction for multiple testing, SSRI-cases exhibited alterations in metabolites related to antioxidant defense such as reduced and oxidized glutathione (GSH and GSSG, respectively), cysteinylglycine, dehydroascorbic acid (DHA), and biliverdin (Figure 2). Furthermore, SSRI-cases exhibited increased levels of serylleucine, citric acid, and monoacylglycerides (1-monostearin, 1-monopalmitin) and decreased levels of nicotinamide adenine dinucleotide (NADH), and phosphocreatine, as well as metabolites possibly linked to external exposure, epigallocatechin and myristamine oxide. However, the 11 metabolites with biological relevance were chosen for further investigation via linear regression analysis, where the impact of SSRI usage remained significant for all metabolites (Supplementary Table 4). Moreover, biliverdin and cysteinylglycine levels were influenced by the mode of delivery, and GSH and cysteinylglycine levels were impacted by preeclampsia.

**Figure 2.**
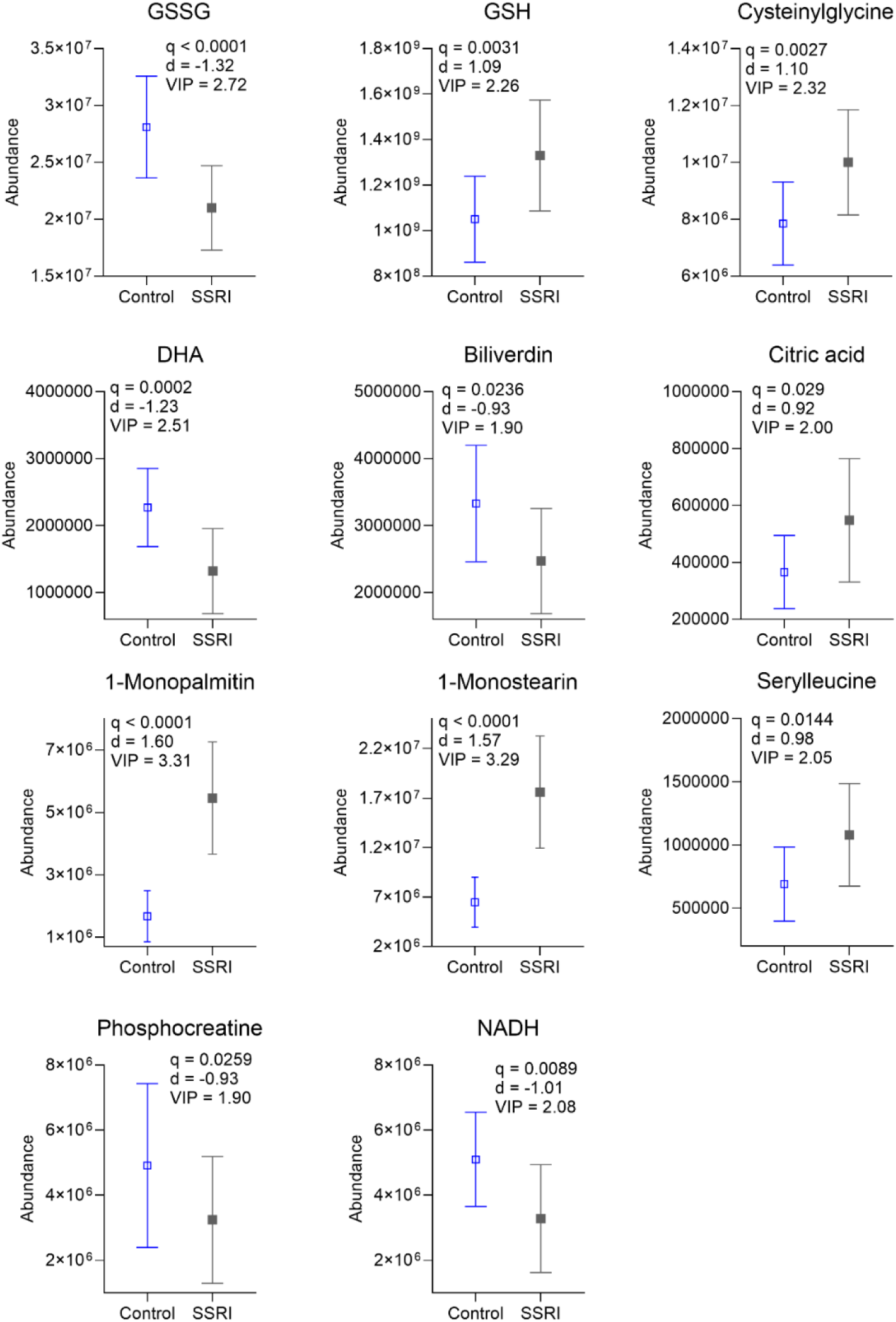
The identified and significantly altered metabolites with biological relevance in selective serotonin reuptake inhibitor (SSRI)-cases (n=24) and controls (n=24). Group means and standard deviations are shown. All the results shown remained significant when confounding lifestyle and pregnancy related variables were considered. GSSG, oxidized glutathione; GSH, reduced glutathione; DHA, dehydroascorbic acid; NADH, nicotinamide adenine dinucleotide; q, FDR corrected p-value; d, Cohen’s d effect size; VIP, Variable Importance in Projection value from partial least sum of squares discriminant analysis

Molecular features with p-value <0.05 and q-value >0.05 were considered as trends. Increasing trends among SSRI-cases were observed e.g., in the levels of several leucine dipeptides, as well as adenosine monophosphate, guanosine-5’-monophosphate, 1-methyl adenosine, and gamma-glutamyl-cysteine, whereas decreasing trends were observed among SSRI-cases e.g., in the levels of several phosphatidylcholines, adenosine diphosphate, 3-indolepropionic acid, cholesterol 7-ketocholesterol, and N-stearoyl-D-sphingomyelin (Figure 1C, Supplementary Table 2).

### Metabolites and maternal-neonatal traits

Among the newborns born to SSRI-cases, the Apgar score at 1 minute positively correlated with GSH (r = 0.46, p = 0.02), cysteinylglycine (r = 0.44, p = 0.03), and citric acid (r = 0.44, p = 0.03) (Figure 3). The Apgar score at 5 minutes positively correlated with GSH (r = 0.55, p = 0.01), cysteinylglycine (r = 0.57, p = 0.004), citric acid (r=0.50, p=0.01), and 1-monostearin (r=0.44, p=0.03). Moreover, newborn birth weight positively correlated with NADH (r = 0.42, p = 0.04). Interestingly, in the control group, the effect was the opposite, yet insignificant. We found no associations between placental weight and significant metabolites.

**Figure 3.**
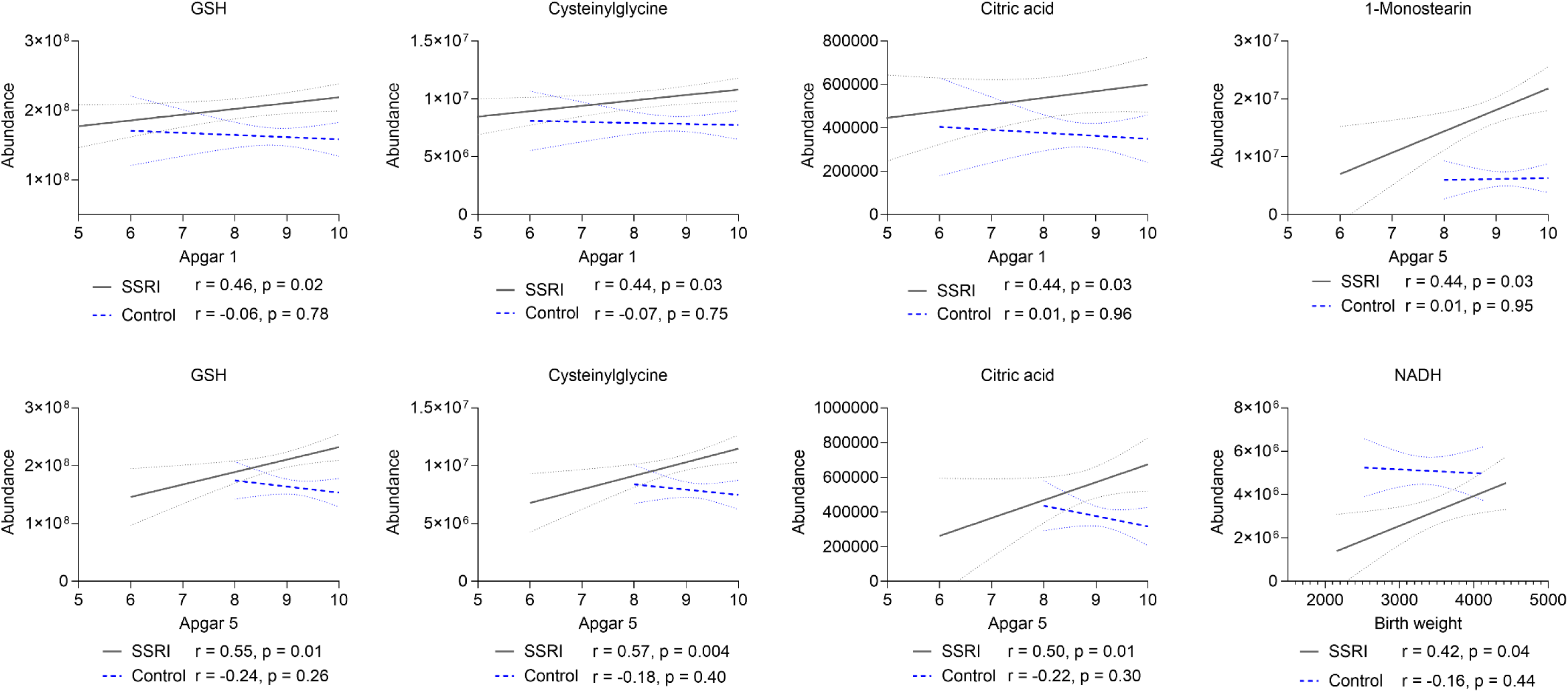
Spearman’s correlation coefficient (r) shows the significant associations between the measured variables and Apgar scores at 1 and 5 minutes and newborn weight among selective serotonin reuptake inhibitor (SSRI) -cases (n = 24) and controls (n = 24). GSH, reduced glutathione; NADH, nicotinamide adenine dinucleotide

### Calculated indicators of placental dysfunction and toxicity

The placental weight and birth weight ratio is used as a marker of placental efficiency and fetal growth [15]. Furthermore, the ratio of GSH to GSSG (GSH:GSSG ratio) is a marker of oxidative stress, an increased ratio implicating enhanced antioxidant activity [16]. Therefore, we explored the two ratios in our cohort. Considering the placental weight and birth weight ratio, we found no significant differences between the groups (Table 1). However, SSRI-cases had nearly twofold GSH:GSSG ratio (M=10.02 vs. 5.92, p<0.001, d=2.24) compared to controls and the impact of SSRI usage remained significant after adjusting for confounding lifestyle and pregnancy related variables (Supplementary Table 4), reflecting efficient glutathione recycling.

## DISCUSSION

The present study aimed to characterize differences in the placental metabolome between SSRI-treated pregnancies and controls. To our knowledge, this is the first study to examine the impact of SSRI use during pregnancy on the human placental metabolome. We detected significant alterations in the metabolite profiles suggesting a potential response to modified oxidative stress status and energy metabolism within the placentas from the SSRI-treated pregnancies.

Along with other mechanisms, glutathione works as the principal non-enzymatic defense mechanism against accumulation of reactive oxygen species (ROS) [17]. In our cohort, glutathione metabolism was notably altered, with increased GSH (d = 1.09), decreased GSSG (d = –1.32), and an elevated GSH:GSSG ratio (d = 2.24). SSRI-cases also showed increased cysteinylglycine (d = 1.10), a glutathione breakdown product, and a trend toward increased gamma-glutamyl-cysteine, a precursor to glutathione, suggesting upregulation of glutathione synthesis. The observed findings align with Ferreira et al. (2020) who suggested that enhanced antioxidant defense in preeclamptic placentas may represent a protective adaptation against oxidative stress [18].

Ascorbic acid metabolism, which shares a pathway with glutathione [19], also showed changes. SSRI-cases had decreased level of DHA (d = –1.23), the oxidized form of ascorbic acid, and a trend toward increased ascorbic acid, suggesting enhanced recycling of these antioxidants. Furthermore, the abundance level of another antioxidant, biliverdin, a product of heme metabolism, was decreased in SSRI-cases (d=-0.93). Oxidative stress activates heme oxygenase 1, which converts heme to biliverdin [20]. Biliverdin is rapidly reduced to bilirubin under physiological conditions [21]. However, we observed no differences in bilirubin levels between groups.

Overall, the alterations in glutathione, ascorbic acid, and biliverdin metabolism indicate enhancement of the antioxidative defense system. The placenta generates both pro-oxidant agents such as ROS and antioxidants, maintaining a robust antioxidant defense system [22]. However, intrinsic and environmental factors can disrupt this redox balance, leading to oxidative stress. Oxidative stress modifies cellular proteins, reflecting changes in enzyme activities and normal cell functions [23]. Redox reactions are tightly coupled with energy metabolism, particularly adenosine triphosphate (ATP) availability [24]. Mitochondrial ROS disrupt most cellular metabolic processes, such as the citric acid cycle, urea cycle, amino acid metabolism, heme synthesis, and fatty acid oxidation [25]. Consequently, the alterations in redox homeostasis could be potentially linked with the observed changes in energy metabolism.

Notably, the level of citric acid, a key metabolite in the tricarboxylic acid (TCA) cycle [26], was elevated in SSRI-cases (d = 0.92). In contrast, NADH, another crucial metabolite involved not only in the TCA cycle but also in glycolysis and fatty acid oxidation [27], was reduced in SSRI cases (d = -1.01). Additionally, we observed alterations in the levels of phosphocreatine (d = -0.93), monoacylglycerides, including 1-monopalmitin (d = 1.60) and 1-monostearin (d = 1.57) and dipeptide serylleucine (d=0.98).

Phosphocreatine plays a crucial role in maintaining ATP availability [28] and creatine metabolism might influence placental bioenergetics and fetal development [29]. Hypoxic placentas rely more on creatine metabolism to stabilize ATP levels, manifesting as increased system function [30]. Our findings of decreased phosphocreatine may suggest reduced local creatine synthesis or altered energy demands, yet the implications remain unclear. Finally, monoacylglycerides are part of the glycerolipid/free fatty acid cycle, which regulates energy expenditure and cell signaling [31], whereas dipeptides serve as a source of amino acids for the developing fetus, supporting protein synthesis [32].

Oxidative stress has been linked to pathogenesis of depression [33]. On the contrary, SSRIs alleviate oxidative stress in unipolar and antenatal depressive patients [34,35] and reverse the reduction in circulating ascorbic acid levels caused by depression in non-pregnant individuals [36]. Indeed, SSRIs impact redox homeostasis, possessing both pro- and antioxidant effects depending on the redox state, tissue, and dose [37]. In meta-analysis of observational studies, SSRIs were suggested to serve as antioxidants [38] and 5-HT itself possesses antioxidant effects in vitro [39]. Jorgensen et al. (2022) proposed that inhibiting SERT raises extracellular 5-HT in the periphery or central nervous system potentially reducing intracellular oxidative stress by altering mitochondrial function via 5-HT_2A_ (5-HT receptor) activation [35]. Arora et al. (2025, preprint) demonstrated that sertraline and paroxetine reduced both ROS and ATP levels in neuroepithelial stem cells, indicating potential mitochondrial impairment [40]. In rodents, SSRIs appear to alleviate oxidative changes in the presence of pre-existing oxidative insult by targeting both enzymatic and non-enzymatic defense mechanisms, such as increasing GSH levels [37]. Moreover, rodent studies indicate that SSRIs may have diverse effects on the antioxidant enzyme glutathione-S-transferase (GST), ranging from activation to inhibition, or showing no significant impact [37], whereas studies in human placental GST enzyme suggest an inhibitory effect of a specific SSRI, fluoxetine [12].

The findings may point to underlying pathophysiological mechanisms of depression and related disorders, which SSRIs might influence through modulation of redox homeostasis. Regardless, the observed changes could contribute to pregnancy complications or influence fetal programming, leading to long-term effects on the fetus. The placental homeostasis of antioxidants, amino acids, and lipids is crucial for optimal fetal development and pregnancy outcomes [41,42]. These components play significant roles in mitigating oxidative stress, ensuring nutrient supply, and regulating fetal growth. Thus, impairments of these processes may lead to adverse pregnancy outcomes and long-term health issues for the child. ROS serve as signaling molecules during neurodevelopment, and their reduction during critical developmental windows may hinder neuronal growth [43]. Placental oxidative stress is linked to pregnancy complications such as gestational diabetes mellitus (GDM), preeclampsia, and intrauterine growth restriction (IUGR), characterized by trophoblast apoptosis and deportation, and changes in placental vascular reactivity [44]. Consequently, it is essential to thoroughly characterize the molecular pathways through which SSRIs potentially modulate placental redox homeostasis. In our cohort, GDM was not associated with the metabolites, whereas preeclampsia significantly altered the associations between SSRI use and both GSH and cysteinylglycine levels. However, the small sample size (n = 3) of preeclamptic placentas raises questions about the validity of the effect.

Furthermore, the clinical significance of observed associations with newborn wellbeing requires further investigation. Notably, GSH, cysteinylglycine, citric acid and 1-monostearin, showed a positive correlation with the 1- and 5-minute Apgar score in SSRI-cases, suggesting that higher levels of these metabolites may be associated with better birth outcomes.

A strength of the current study is the possibility of assaying metabolites directly in placental tissue since commonly in clinical studies determination of circulatory or urinary levels is possible due to ethical limitations. According to the clinical records, the cases consistently used SSRIs throughout their pregnancy, minimizing the risk of errors due to medication discontinuation.

Our study has potential limitations. Although the largest to date, the study sample is relatively small. Other limitations include the inability to distinguish the effects of SSRIs from those of mental health disorders, as well as to measure the treatment response to SSRIs. Furthermore, metabolite annotation remains as a major limitation in metabolomics studies, particularly in less studied organs such as the placenta. Given the limited research on placental metabolomics, we believe the findings of this exploratory study are significant. However, the validation of the findings, targeted analysis of selected metabolites such as GSH and GSSG remains to be a subject of future studies.

In conclusion, this study provides important new insights into SSRI-induced alterations in the human placenta and, to our knowledge, is the first to investigate impact of SSRIs on the human placental metabolome during pregnancy. The findings suggest a complex metabolic shift, potentially indicating a response to changes in redox homeostasis and energy metabolism. Furthermore, specific metabolites could potentially reflect birth outcomes. Since the placenta is a metabolically active organ and fetal development heavily depends on its proper function, further studies are needed to elucidate the clinical significance of the observed effects.

## MATERIALS AND METHODS

### Ethics

All participants in the Kuopio Birth Cohort (KuBiCo) study provided a written informed consent. The study was conducted in accordance with the guidelines outlined in the Declaration of Helsinki. Ethical approval was obtained from the Research Ethics Committee of the Hospital District of Central Finland in Jyväskylä, Finland on November 15, 2011 (18U/2011). The study was conducted in accordance with the Strengthening the reporting of observational studies in epidemiology (STROBE) guidelines.

### Clinical subjects and sample collection

A total of 48 placentas from individuals using SSRI medication throughout the pregnancy (n=24) and non-depressive control individuals without antidepressant medication (n=24) were included in the study. The placental sample collection is standardized within the KuBiCo [45]. The samples originate from pregnant individuals donating placenta after giving birth at Kuopio University Hospital (KUH) during the years 2013-2015. The samples were collected by midwives in the delivery room. Twenty pieces from the central area of placenta containing basal plate and villi were collected after delivery, divided into four tubes, frozen in liquid nitrogen, and stored at -80°C.

Data on SSRI use and the diagnosis of GDM (ICD-10 code O24.4) and preeclampsia (ICD-10 code O14-O15) were collected from the KUH Birth Register, which contains demographic and clinical information about the study participants. The Birth register also included a self-reporting section to report the use of over-the-counter and prescription drugs, as well as smoking during pregnancy. The smoking status of participants was confirmed by measuring serum cotinine level from blood samples collected at birth using a previously described method [11].

### Sample preparation

Prior to LC-MS analysis, placental samples were subjected to cryogenic grinding to homogenize the tissue. Approximately 1-2 grams of the deep frozen (-80°C) tissue was grinded with QIAGEN^®^ TissueLyser II for one minute (Frequency 30.0 1/s). Powdered frozen tissue was stored in -80°C until the metabolomics analysis.

### Nontargeted LC-MS profiling

The samples were analyzed with two LC-MS methods [46]. Prior to analysis, 80% methanol was added to the powdered placental tissue at a ratio of 1:3 (e.g.,100 mg of sample + 300 µl of 80% methanol). The samples were carefully mixed and centrifuged for 10 minutes at +4°C and 16100 rcf. The samples were then diluted 1:1 with 80% methanol (200 µl of sample + 200 µl of 80% methanol). The diluted samples were filtered using a syringe filter (15161499, PTFE syringe filter 13mm, 0.2 µm) into a high-performance liquid chromatography vial insert. For more hydrophilic compounds, hydrophilic interaction chromatographic technique (HILIC) was used with both positive and negative electrospray ionization. For more lipophilic compounds, we utilized reversed phase (RP) chromatography and collected data with both electrospray ionization polarities. Both chromatographic techniques were connected to high-accuracy and high-resolution mass spectrometer. A pooled sample from all placental samples was injected at the beginning and end of the analysis to equilibrate the analytical platform and then after every 12 samples throughout the analysis for quality control.

We employed the open-source software MS-DIAL version 4.9.221218 for peak picking, peak alignment, and metabolite identification [47]. Consequently, we conducted preprocessing of the metabolomics data employing “notame” R-package (version 0.3) [48]. Key preprocessing steps involved drift correction, flagging low-quality features, and missing value imputation. The data quality is monitored at each step of the preprocessing according to the protocol detailed in Klåvus et al. (2020) [48].

### Statistical analysis

For the clinical characteristics, continuous variables were analyzed with Welch’s t-test or Wilcoxon rank-sum test for parametric and nonparametric variables, respectively. Categorical variables were analyzed with Fisher’s exact test.

For feature-wise analysis of the metabolomics data, we employed Welch’s t-test to calculate p-values for each molecular feature independently, employing FDR-correction to address the issue of multiple testing. Q-values (FDR corrected p-values) <0.05 were considered statistically significant, whereas molecular features with p-value <0.05 but q-value >0.05 were considered trends. Moreover, Cohen’s d effect sizes were computed to measure the magnitude of differences between the study groups. A positive Cohen’s d value indicates that the metabolite abundance was increased in SSRI-cases, and a negative value indicates a decrease in SSRI-cases compared to controls. For multivariate analysis of the metabolomics data, we used principal component analysis (PCA), and partial least sum of squares discriminant analysis (PLS-DA) calculated with SIMCA (version 17, Sartorius Stedim Data Analytics AB).

To account for potential confounding factors, we conducted a linear regression analysis on metabolites with biological relevance that exhibited statistically significant differences between the SSRI-cases and controls. The initial model (model 1) included merely the group (SSRI vs. control). The selected covariates for modeling possible influence of lifestyle included BMI, cotinine, and co-medication (model 2) and for modeling possible influence of pregnancy we included mode of delivery, fetal sex, preeclampsia, and GDM (model 3) due to their known influences on the maternal metabolome [49–52]. Regression analyses were performed in R Studio (version 4.3.3). Finally, we investigated the associations between the significantly altered metabolites and placental weight, infants’ birth weight and Apgar scores with Spearman’s correlation coefficients.

### Metabolite identification

Following Welch’s t-test, molecular features displaying an FDR-corrected q-value <0.05 underwent metabolite identification, while those with non-corrected p-values <0.05 were also analyzed for trends. The identified metabolites were categorized into identification levels 1 and 2 according to the community standards [53].

## ACKNOWLEDGEMENTS

This paper belongs to the studies carried out by the Kuopio Birth Cohort consortium and we thank our colleagues who are responsible for the design and conduct of the KuBiCo. We thank Satu Marttila, Sanna Koivisto, Miia Reponen, and Juulia Kuparinen for expert laboratory assistance, and the staff of the Department of Obstetrics and Gynecology in Kuopio University Hospital. The authors also want to thank the Biocenter Finland and the Biocenter Kuopio for supporting the core LC-MS laboratory facility. This project has received funding from the European Union’s Horizon 2020 research and innovation programme under grant agreement No 825762.

## CRediT Authorship contribution statement

**Anna Itkonen:** Conceptualization, Formal analysis, Visualization, Data curation, Writing – original draft, Writing – review & editing, **Olli Kärkkäinen:** Conceptualization, Methodology, Formal analysis, Writing – review & editing, **Marko Lehtonen:** Methodology, Investigation, Writing – review & editing: **Heidi Sahlman:** Conceptualization, Resources, Data curation, Supervision, Writing – review & editing, **Leea Keski-Nisula:** Resources, Data curation, Writing – review & editing, **Jaana Rysä:** Conceptualization, Resources, Supervision, Project administration, Funding acquisition, Writing – review & editing.

## DATA AVAILABILITY

All the data necessary to evaluate the conclusions is included in the paper and the Supporting Information. The authors are willing to provide additional data related to this paper upon reasonable request.

