## Supplementary Table 1 for "METABOLOMIC INSIGHTS: LC-MS PROFILING OF HUMAN PLACENTAL TISSUE FROM SSRI-TREATED PREGNANCIES"

**Table 1.** The dosage and indications for selective serotonin reuptake inhibitors (SSRIs).

| **SSRI** | **Dose (mg)** | **n (%)** |
|---|---|---|
| Citalopram | 5–40 | 12 (50.0) |
| Escitalopram | 10–20 | 6 (25.0) |
| Sertraline | 25–100 | 5 (20.8) |
| Fluoxetine | 60 | 1 (4.2) |
| **Indications for SSRIs** | | |
| Depression | | 15 (62.5) |
| Panic disorder | | 5 (20.8) |
| Anxiety | | 2 (8.3) |
| Bipolar disorder | | 1 (4.2) |
| Muscle spasm | | 1 (4.2) |

Data is shown as n (%).
