## Supplementary Table 4 for "METABOLOMIC INSIGHTS: LC-MS PROFILING OF HUMAN PLACENTAL TISSUE FROM SSRI-TREATED PREGNANCIES"

**Supplementary Table 3** Linear regression models with the metabolite as the dependent variable and (model 1) group (SSRI vs. control) as a cofactor. For modeling possible influence of lifestyle (model 2) we included body mass index at delivery (BMI), cotinine, and co-medication (model 2) as cofactors, and for modeling possible influence of pregnancy (model 3) mode of delivery, fetal sex, preeclampsia, and gestational diabetes mellitus (GDM) were included as cofactors.

**Model 1: Adjusted for group (SSRI vs. control)**

| | Adj. R2 | F | Beta | p-value |
|---|---|---|---|---|
| **GSH** | 0.30 | 21.12 | 42857205 | **<0.001** |
| **GSSG** | 0.43 | 36.41 | -7161142 | **<0.001** |
| **GSH:GSSG** | 0.56 | 60.21 | 4.10 | **<0.001** |
| **Citric acid** | 0.20 | 12.54 | 182280 | **<0.001** |
| **NADH** | 0.20 | 12.54 | -1813747 | **<0.001** |
| **Cysteinylglycine** | 0.25 | 16.28 | 2171837 | **<0.001** |
| **Serylleucine** | 0.23 | 14.79 | 393377 | **<0.001** |
| **Phosphocreatine** | 0.20 | 12.91 | -1018388 | **<0.001** |
| **Biliverdin** | 0.20 | 12.91 | -859861 | **<0.001** |
| **DHA** | 0.37 | 28.78 | -946421 | **<0.001** |
| **1-Monopalmitin** | 0.65 | 88.33 | 3795111 | **<0.001** |
| **1-Monostearin** | 0.62 | 77.46 | 11160717 | **<0.001** |

**Model 2: Adjusted for group (SSRI vs. control), BMI, cotinine, co-medication**

| | Adj. R2 | F | Beta | | | | p-value | | | |
|---|---|---|---|---|---|---|---|---|---|---|
| | | | SSRI usage | BMI | Cotinine | Co-medication | SSRI usage | BMI | Cotinine | Co-medication |
| **GSH** | 0.29 | 5.77 | 40227754 | -401477 | -474949 | -5148275 | **<0.001** | 0.576 | 0.128 | 0.593 |
| **GSSG** | 0.40 | 8.71 | -7193732 | -38219 | 32383 | -174395 | **<0.001** | 0.700 | 0.448 | 0.896 |
| **GSH:GSSG** | 0.55 | 15.20 | 4 | 0 | 0 | 0 | **<0.001** | 0.851 | 0.069 | 0.870 |
| **Citric acid** | 0.18 | 3.58 | 173894 | -314 | -2927 | -18497 | **0.004** | 0.940 | 0.107 | 0.741 |
| **NADH** | 0.22 | 4.29 | -1884479 | -28543 | 6715 | -538417 | **<0.001** | 0.443 | 0.674 | 0.283 |
| **Cysteinylglycine** | 0.30 | 5.82 | 2011629 | -11004 | -28860 | -323884 | **<0.001** | 0.768 | 0.077 | 0.518 |
| **Serylleucine** | 0.22 | 4.27 | 371767 | 4980 | -6001 | 69891 | **0.002** | 0.530 | 0.083 | 0.511 |
| **Phosphocreatine** | 0.17 | 3.30 | -914087 | -21499 | 7439 | 80029 | **0.008** | 0.363 | 0.464 | 0.800 |
| **Biliverdin** | 0.14 | 2.91 | -884075 | -664 | 4059 | -132111 | **0.003** | 0.974 | 0.638 | 0.623 |
| **DHA** | 0.41 | 8.95 | -918735 | -21726 | -7551 | -231467 | **<0.001** | 0.121 | 0.208 | 0.216 |
| **1-Monopalmitin** | 0.64 | 21.41 | 3604968 | -4978 | 17842 | -485108 | **<0.001** | 0.880 | 0.211 | 0.275 |
| **1-Monostearin** | 0.61 | 18.82 | 10783478 | -62287 | 25684 | -1844369 | **<0.001** | 0.550 | 0.567 | 0.191 |

**Model 3: Adjusted for group (SSRI vs. control), mode of delivery, fetal sex, preeclampsia, GDM**

| | Adj. R2 | F | Beta | | | | | p-value | | | | |
|---|---|---|---|---|---|---|---|---|---|---|---|---|
| | | | SSRI usage | Mode of delivery | Fetal sex | Preeclampsia | GDM | SSRI usage | Mode of delivery | Fetal sex | Preeclampsia | GDM |
| **GSH** | 0.52 | 10.61 | 50651319 | -24760738 | -5284146 | -49302765 | 8205384 | **<0.001** | 0.072 | 0.587 | **0.009** | 0.479 |
| **GSSG** | 0.41 | 7.16 | -6591687 | -1406921 | 1412770 | -489138 | -231900 | **<0.001** | 0.497 | 0.345 | 0.859 | 0.896 |
| **GSH:GSSG** | 0.57 | 12.69 | 4.28 | -0.79 | -0.60 | -1.67 | 0.07 | **<0.001** | 0.396 | 0.371 | 0.183 | 0.934 |
| **Citric acid** | 0.35 | 5.73 | 226726 | -111724 | 2762 | -167373 | 46342 | **<0.001** | 0.158 | 0.961 | 0.115 | 0.491 |
| **NADH** | 0.25 | 3.91 | -1622158 | -279963 | 897514 | -285967 | -509152 | **0.003** | 0.721 | 0.118 | 0.784 | 0.450 |
| **Cysteinylglycine** | 0.55 | 11.90 | 2632702 | -1552938 | -271502 | -2380127 | 493702 | **<0.001** | **0.024** | 0.572 | **0.010** | 0.389 |
| **Serylleucine** | 0.18 | 2.94 | 437579 | -75667 | 8292 | -129617 | -136056 | **0.001** | 0.687 | 0.951 | 0.606 | 0.401 |
| **Phosphocreatine** | 0.23 | 3.58 | -1102010 | 71614 | -358113 | -323604 | -417737 | **0.001** | 0.883 | 0.312 | 0.620 | 0.321 |
| **Biliverdin** | 0.24 | 3.79 | -806992 | 883542 | 479236 | -503457 | -212581 | **0.003** | **0.031** | 0.100 | 0.345 | 0.533 |
| **DHA** | 0.33 | 5.30 | -922370 | -224047 | -134361 | -303697 | 67141 | **<0.001** | 0.486 | 0.562 | 0.479 | 0.807 |
| **1-Monopalmitin** | 0.70 | 21.68 | 4050668 | 145883 | 507588 | -1304645 | 253852 | **<0.001** | 0.823 | 0.285 | 0.140 | 0.651 |
| **1-Monostearin** | 0.69 | 20.87 | 11998934 | 126388 | 2443123 | -4667196 | 292869 | **<0.001** | 0.948 | 0.089 | 0.080 | 0.861 |

SSRI, selective serotonin reuptake inhibitor; GSH, reduced glutathione; GSSG, oxidized glutathione; NADH, nicotinamide adenine dinucleotide; DHA, dehydroascorbic acid
